# IMPROVING THE MANAGEMENT OF LEPROSY ULCERS THROUGH A COMMUNITY SELF-CARE INTERVENTION USING A STEPPED WEDGE CLUSTER RANDOMISED TRIAL IN SOUTH-EAST NIGERIA

**DOI:** 10.1101/2025.02.10.25321982

**Authors:** Anthony O Meka, Linda Ugwu, Yixin Wang, Samuel I. Watson, Onaedo Ilozumba, Uchechukwu Akunna, Sopna Choudhury, Joseph N Chukwu, Paramjit Gill, Antje Lindenmeyer, Ngozi Murphy-Okpala, Nchekwube Ndubuizu, Jo Sartori, Frances Griffiths, Richard J. Lilford

## Abstract

**Background:** People with sensory neuropathy caused by leprosy suffer from recurrent ulcers. Self-care is used to promote ulcer healing and reduce recurrence. The challenge is to maximise use of self-care and to optimise the way it is practised. RedAid Nigeria in South-East Nigeria adapted and implemented a recently developed international guideline for self-care of ulcers in an area with a high prevalence of leprosy. We report a prospective evaluation of the effectiveness of this intervention in reducing the prevalence and severity of ulcers and in improving wellbeing.

**Methods:** A mixed methods partial stepped wedge cluster randomized trial across 15 clinics serving patients affected by neuropathy and recurrent ulcers. Outcomes were number and area of ulcers, self-rated health and wellbeing. We used a Bayesian analysis to estimate treatment effects and a process analysis based on descriptive statistics and thematic analysis of qualitative data.

**Results:** The ulcers at baseline were long-standing – mean (sd) of ulcer duration at baseline was 58.55 (94.49) months and total disability rate was high (65.99%). There was no evidence of an intervention effect on the persistence of ulcers (OR: 1.00 (95% credible interval (CrI) 0.29 – 3.56) or relative area of ulcers (cm^2^) 1.12 (0.53 – 2.27). There was evidence of an increase in self-reported health and wellbeing associated with the intervention (EQ-5D VAS mean difference 10.93 (8.60, 13.23) out of 100; life satisfaction 1.18 (0.65, 1.72) out of 10). Process data indicate the intervention was delivered as planned. Qualitative data triangulated with quantitative data findings suggest enthusiasm among participants for and adherence to the intervention. Improved wellbeing was described in terms of improved confidence in administering self-care and better integration in local communities.

**Discussion:** The intervention improved perceptions of health and wellbeing, despite having small and uncertain effects on ulcer-related outcomes. Effecting an improvement in ulcer size and number through a community intervention may be difficult to achieve in poor, rural contexts, even if patients respond positively to the intervention.

Trial Registration: Retrospectively registered on the ISRCTN registry with trial registration number ISRCTN12572449 and can be viewed at: https://www.isrctn.com/ISRCTN12572449.

## Introduction

Patients affected by leprosy do not commonly present with an ulcer at the time of diagnosis. As in diabetes, ulcers are caused by nerve damage and the resulting loss of sensation. Neuropathy in leprosy is caused primarily by inflammatory ‘reactions’, which occur in 30-70% of leprosy cases in response to live *Mycobacterium leprae* or persistent cell wall antigens in skin and nerve tissues (1). Those experiencing recurrent ulcers often face significant consequences, including diminished function, financial setbacks, and social stigma. As a result people suffer chronic depression and withdrawal from society (2). The standard method for both treatment and prevention of neuropathic ulcers is a programme of “self-care”. The World Health Organization (WHO) defines self-care as ‘the ability of individuals, families and communities to promote health, prevent disease, maintain health, and cope with illness and disability with or without the support of a health-care provider’ (3). The aim of self-care is to enable affected people to manage their diseases and continue with their normal activities. Self-care in leprosy follows a sequence, beginning with visual inspection of the skin and limbs to identify early stages of ulcer formation followed by intensive skin care processes including soaking, scraping, and oiling ulcerated or vulnerable surfaces. Above all self-care relies on reducing pressure on affected surfaces by walking less and/or walking with the use of crutches or an off-loading device such as a specifically designed shoe (4, 5).

Self-care is supported by evidence from a recent systematic review which reported that about two-thirds of ulcers in diabetes can be avoided through self-care (6). Similar results have been reported in leprosy in Nigeria (7). Recent guidelines from the WHO and the International Federation of Anti-leprosy Associations (ILEP) strongly recommend self-care. However, self-care does not exist in a vacuum but to be sustainable it must be supported by the broader system in which it is located (8). The WHO Global Leprosy Strategy has identified the need for “*research to build models that demonstrate how participation of affected persons can be structurally embedded in various contexts”* and “*research into involvement of different community groups*”(9). Building on these principles, the NIHR Research and Innovation for Global Health Transformation programme funded this study which established an international consensus programme to develop a set of guidelines for self-care (10–13). Consistent with the above WHO principles, these guidelines not only describe optimal self-care itself, but also the mechanisms to support the front line of care, including “train the trainers” programmes.

The study reported here is the evaluation of implementation of the above guidelines to provide a self-care intervention. The intervention was implemented, and part funded by, RedAid Nigeria (formally German Leprosy and TB Relief Association-DAHW) situated in South-East Nigeria. Support from RedAid Nigeria thus represents intensification of an existing programme in line with the above guidelines. We hypothesised that an intervention built on organisational and behavioural principles combined with clinical support can break the cycle of repeated ulceration in leprosy. We hypothesised further that wellbeing would be improved.

## Methods

### Study design

A “staircase” of incomplete stepped-wedge cluster randomized design (14) with masked / blinded quantitative outcomes and qualitative observations. The protocol for the intervention is available online (15). We report this study according to CONSORT guidelines for stepped-wedge trials (16).

### Participants

#### Study sites

The study was conducted in South-East Nigeria in the vicinity of 2 hospitals each serving communities with a high prevalence of leprosy (Mile Four Hospital, Abakaliki, Ebonyi State and St. Benedict’s Hospital Ogoja in Cross River State, Figure 1). Mile Four hospital serves ten outstation clinics and St Benedict’s Hospital serves five outstation clinics. Each outstation clinic is situated in a village. Each village supported one self-care group. These groups form the clusters for the statistical analysis. However, the process evaluation, while focusing mostly on the self-care groups, nevertheless went broader than the group itself to include the community within each village.

**Figure 1:**
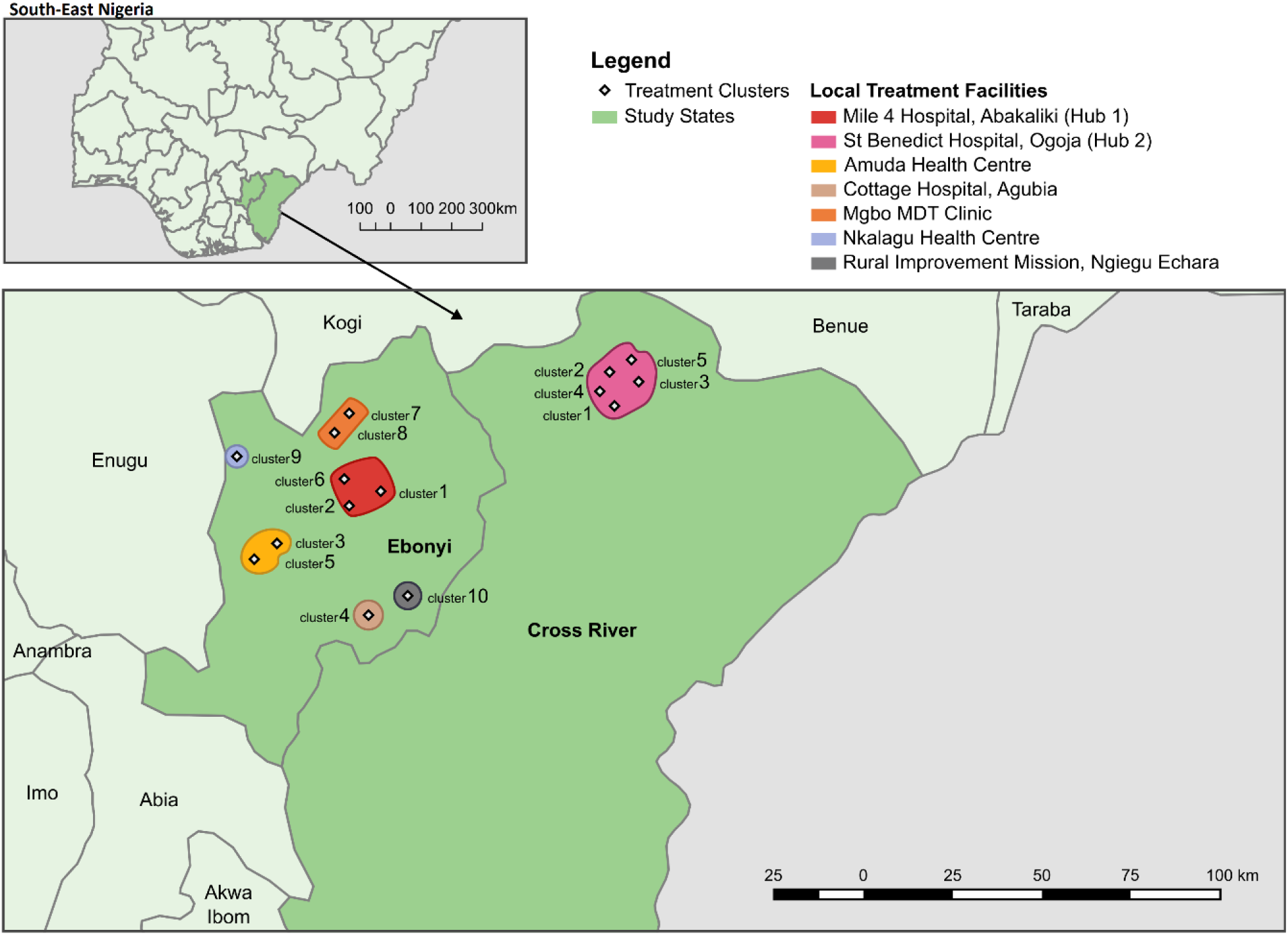
cluster locations within 2 participating states.

#### Individual participants

Eligible patients were identified by the implementation team (see below) using registers of people at risk of leprosy ulcers held in the two study centre hospitals. Participants were people affected by leprosy/with a high risk of limb ulceration based on an existing ulcer, history of ulcer, or sensory loss in one or more extremity. Potential participants were informed about the study by health workers employed by local government and health care providers based at the above hospitals. Participants were invited to take part in the trial by a research fellow trained to Good Clinical Practice standards. A patient information leaflet was given to potential participants, and they were asked to sign a consent form. The patient information leaflet and consent forms were translated from English and then back translated using the WHO methodology for accurate translation (17). If patients agreed to take part, they were recruited at baseline before the clusters were randomized to a designated period in the intervention sequence. Although data collection and intervention staff were employed by RedAid, the data collection staff were independent of the implementation team. Participants were recruited between April 2021 and August 2022.

### Randomisation and masking

To support implementation, the clusters were grouped according to hospital catchment area. The 15 clusters were randomly allocated into one of five sequences (steps). The randomization was restricted geographically to facilitate the logistics of implementation in the Mile Four hospital catchment area. Thus, within the Mile Four hospital area, the ten clusters were grouped into five geographically proximal units, which were then randomly allocated to one of five trial sequences. The remaining five clusters, serving St. Benedict’s hospital, were separately randomly allocated to the five trial sequences. Figure 2 shows the trial sequences and cluster allocations. The randomization process was carried out by the trial statistician at the University of Birmingham.

**Figure 2:**
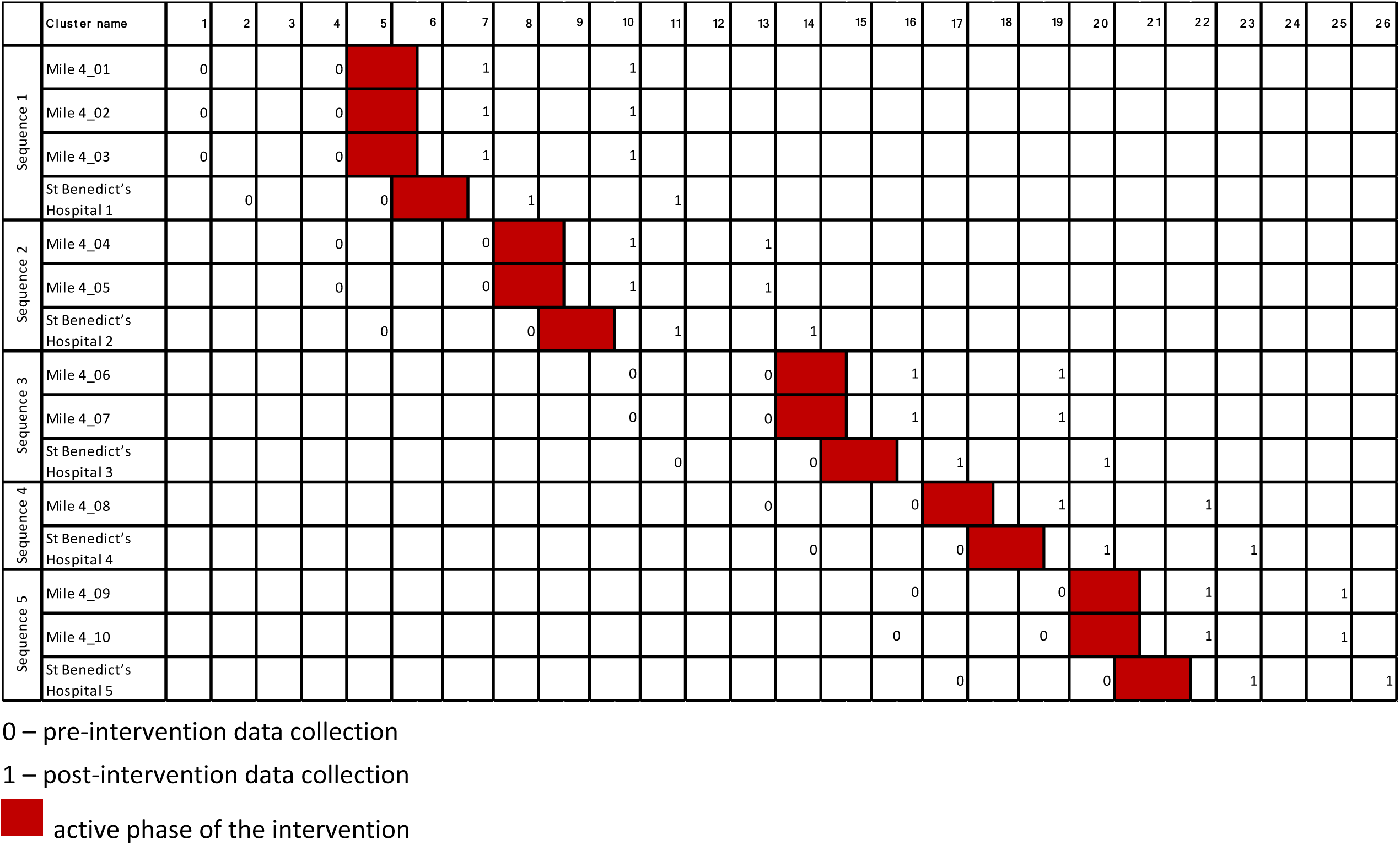
Sequencing of the intervention for months 1-26. Each row represents a cluster.

### The intervention

As stated, the intervention was an evidence informed intervention, based on the Consensus Guidelines for Self-Care Interventions for Leprosy developed by an International Scientific Advisory Committee. The Committee included public, community and policy representatives, as well as academics and researchers experienced in implementation science. ‘The Guidelines’ were informed by psychological and organisational theory (10, 13, 18–20) and based on the current state of science. The guidelines have been published elsewhere (10–13). However, given the variation in contexts within which leprosy occurs, the guidelines were designed to be adapted for local conditions (21). Here we describe how the guidelines were adapted for use in South-East Nigeria.

RedAid Nigeria worked with different local stakeholder groups to adapt ‘The Guidelines’. The groups comprised expert clinical staff and ulcer researchers (dealing with leprosy and with other chronic conditions), representatives of the International Association of Anti-Leprosy Organisations (ILEP) and the government disease control programme, and people with lived experience of leprosy. An intervention manual was co-produced, detailing the critical aspects of steps in self-care practice in Nigeria and describing how ‘The Guidelines’ were adapted to ensure that they were culturally appropriate and accessible for the setting. Participant-facing materials were co-designed with people affected by leprosy, (including those who were marginalised, had low literacy and had residual effects of leprosy). The adapted intervention was represented in a manual.

A detailed TIDieR compliant description of the intervention is included in Supplementary material (S1). Briefly, the intervention was based on re-activation of self-care groups (SCGs) that had ceased to function or enhanced activity of groups that were still active. The intervention can thus be thought of as an increased ‘dose’ based on the above adapted guideline and delivered under the overall supervision of the organisation responsible for leprosy care in the area.

The management cascade is represented in Figure 3. The intervention, based on the manual, was overseen by two officials – one from each site – Ebony and Cross-River, where respectively Mile Four and St Benedicts Hospitals are based. Practical arrangements were delegated to seven permanent staff from these institutions - we refer to these staff members as ‘Programme co-ordinators’ (N=7). These staff trained and mentored community outreach workers, accompanying them on their initial field visits. The facilitators brought the SCGs into existence, supervised elections of an SCG leader from among the SCG members and taught and encouraged the SCGs in the principles and practice of self-care.

**Figure 3:**
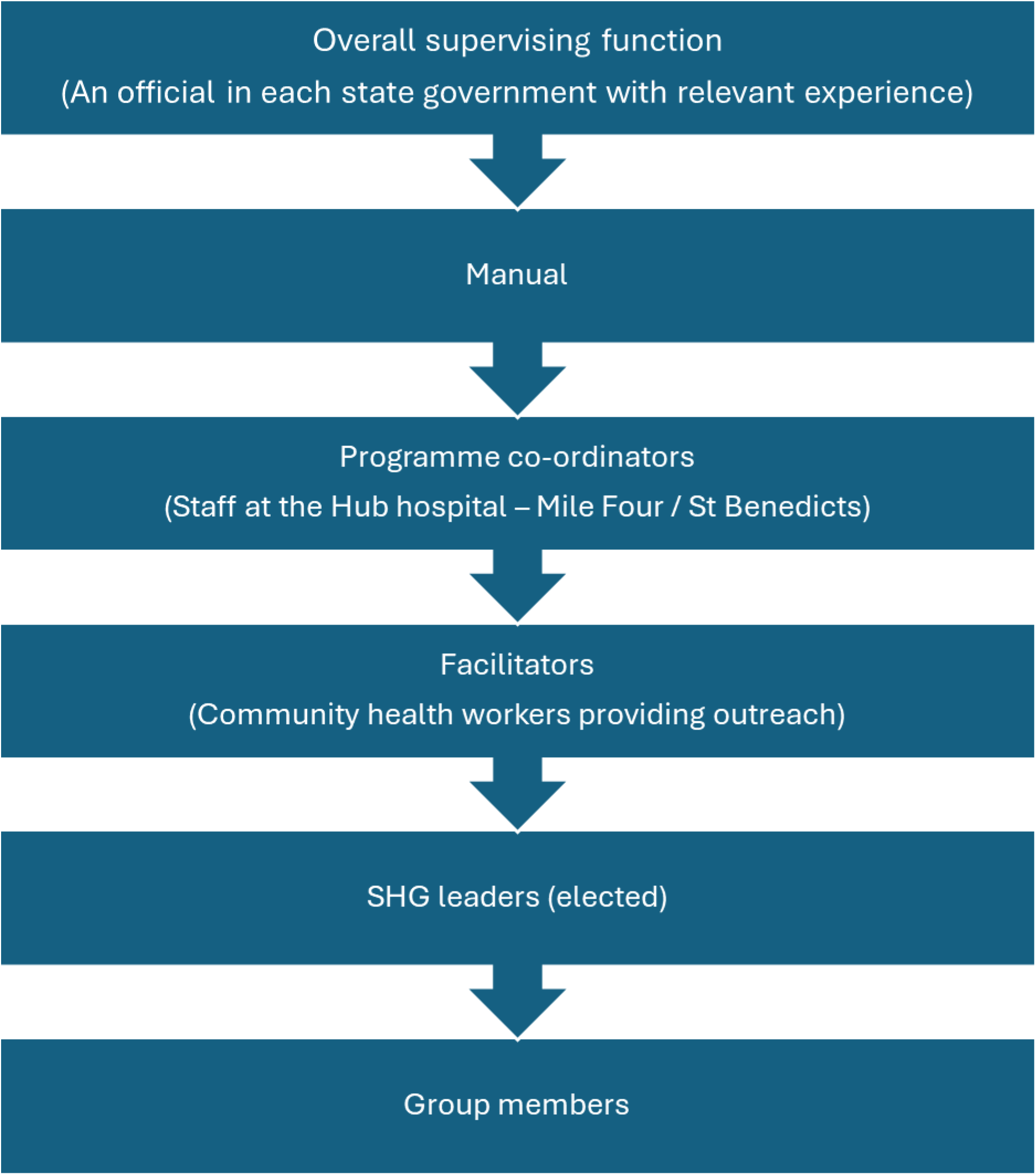
Training cascade to implement the adapted self-care intervention.

The intervention was delivered over a 6 weeks ‘active phase’ (Figure 2). During the active phase, three days of intensive training were delivered at each hub, followed by weekly sessions to re-inform good self-care practice.

The facilitators provided education materials in appropriate formats for use in a low literacy setting.

The six-week ‘active’ phase of the intervention described above was followed by a less intensive ‘sustainability’ phase during which the mentorship visits were reduced to once every six weeks until the end of the intervention on 30^th^ May 2023.

### Data collection

There were four data collection periods. Pre-intervention data was collected on two occasions, three months apart (indicated as zero in Figure 2). Post-intervention data were also collected on two occasions, three months apart and twice after the intervention (indicated as ‘1’ in Figure 2). The research fellows (UA & LU), fluent in the local languages, collected data electronically on Tablets. The questionnaires themselves were in English. No other authors had access to information that could identify individual participants.

Demographic data were collected at baseline i.e. the first interview above (Table 1). Quality of life and well-being questionnaires were administered at all four data collection points (described in detail below). Also, the limbs were inspected at all four data collection points, and their condition described using a standard form with information on 1) peripheral anaesthesia, 2) number of ulcers and 3) any deformities (using the WHO disability grading system (22). The largest ulcer was photographed in a standard manner (23) by the research fellow using the camera in the data collection tablet. A plastic ruler, sterilised in spirit, was placed in the frame at the level of the ulcer. A photograph of that ulcer was repeated over the subsequent three visits (Figure 2). In addition, photographs of the plantar surfaces of the two feet together were taken for quality control purposes (i.e. to cross-reference with data recorded on site).

**Table 1:**
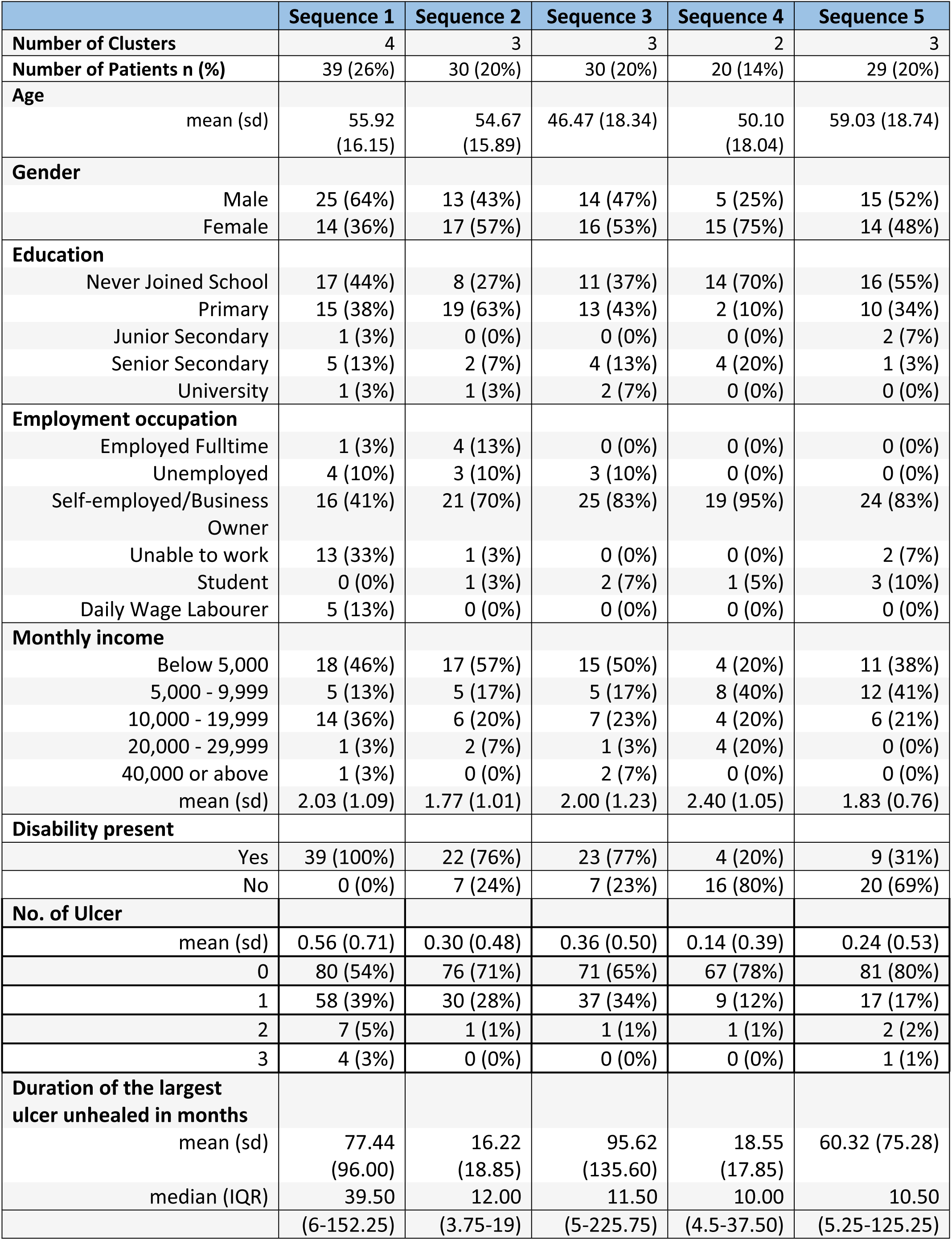
Baseline characteristics.

|  | Sequence 1 | Sequence 2 | Sequence 3 | Sequence 4 | Sequence 5 |
| --- | --- | --- | --- | --- | --- |
| **Number of Clusters** | 4 | 3 | 3 | 2 | 3 |
| **Number of Patients n (%)** | 39 (26%) | 30 (20%) | 30 (20%) | 20 (14%) | 29 (20%) |
| **Age** |  |  |  |  |  |
| mean (sd) | 55.92<br>(16.15) | 54.67<br>(15.89) | 46.47 (18.34) | 50.10<br>(18.04) | 59.03 (18.74) |
| **Gender** |  |  |  |  |  |
| Male | 25 (64%) | 13 (43%) | 14 (47%) | 5 (25%) | 15 (52%) |
| Female | 14 (36%) | 17 (57%) | 16 (53%) | 15 (75%) | 14 (48%) |
| **Education** |  |  |  |  |  |
| Never Joined School | 17 (44%) | 8 (27%) | 11 (37%) | 14 (70%) | 16 (55%) |
| Primary | 15 (38%) | 19 (63%) | 13 (43%) | 2 (10%) | 10 (34%) |
| Junior Secondary | 1 (3%) | 0 (0%) | 0 (0%) | 0 (0%) | 2 (7%) |
| Senior Secondary | 5 (13%) | 2 (7%) | 4 (13%) | 4 (20%) | 1 (3%) |
| University | 1 (3%) | 1 (3%) | 2 (7%) | 0 (0%) | 0 (0%) |
| **Employment occupation** |  |  |  |  |  |
| Employed Fulltime | 1 (3%) | 4 (13%) | 0 (0%) | 0 (0%) | 0 (0%) |
| Unemployed | 4 (10%) | 3 (10%) | 3 (10%) | 0 (0%) | 0 (0%) |
| Self-employed/Business Owner | 16 (41%) | 21 (70%) | 25 (83%) | 19 (95%) | 24 (83%) |
| Unable to work | 13 (33%) | 1 (3%) | 0 (0%) | 0 (0%) | 2 (7%) |
| Student | 0 (0%) | 1 (3%) | 2 (7%) | 1 (5%) | 3 (10%) |
| Daily Wage Labourer | 5 (13%) | 0 (0%) | 0 (0%) | 0 (0%) | 0 (0%) |
| **Monthly income** |  |  |  |  |  |
| Below 5,000 | 18 (46%) | 17 (57%) | 15 (50%) | 4 (20%) | 11 (38%) |
| 5,000 - 9,999 | 5 (13%) | 5 (17%) | 5 (17%) | 8 (40%) | 12 (41%) |
| 10,000 - 19,999 | 14 (36%) | 6 (20%) | 7 (23%) | 4 (20%) | 6 (21%) |
| 20,000 - 29,999 | 1 (3%) | 2 (7%) | 1 (3%) | 4 (20%) | 0 (0%) |
| 40,000 or above | 1 (3%) | 0 (0%) | 2 (7%) | 0 (0%) | 0 (0%) |
| mean (sd) | 2.03 (1.09) | 1.77 (1.01) | 2.00 (1.23) | 2.40 (1.05) | 1.83 (0.76) |
| **Disability present** |  |  |  |  |  |
| Yes | 39 (100%) | 22 (76%) | 23 (77%) | 4 (20%) | 9 (31%) |
| No | 0 (0%) | 7 (24%) | 7 (23%) | 16 (80%) | 20 (69%) |
| **No. of Ulcer** |  |  |  |  |  |
| mean (sd) | 0.56 (0.71) | 0.30 (0.48) | 0.36 (0.50) | 0.14 (0.39) | 0.24 (0.53) |
| 0 | 80 (54%) | 76 (71%) | 71 (65%) | 67 (78%) | 81 (80%) |
| 1 | 58 (39%) | 30 (28%) | 37 (34%) | 9 (12%) | 17 (17%) |
| 2 | 7 (5%) | 1 (1%) | 1 (1%) | 1 (1%) | 2 (2%) |
| 3 | 4 (3%) | 0 (0%) | 0 (0%) | 0 (0%) | 1 (1%) |
| **Duration of the largest ulcer unhealed in months** |  |  |  |  |  |
| mean (sd) | 77.44<br>(96.00) | 16.22<br>(18.85) | 95.62<br>(135.60) | 18.55<br>(17.85) | 60.32 (75.28) |
| median (IQR) | 39.50 | 12.00 | 11.50 | 10.00 | 10.50 |
|  | (6-152.25) | (3.75-19) | (5-225.75) | (4.5-37.50) | (5.25-125.25) |

The ulcer photographs were transferred to the University of Birmingham where the date was removed and the image was given a unique identification number. The image was transferred to the Leprosy Mission Trust India in Delhi, India for independent measurement of the ulcer area. There, local research fellows used the electronic Pressure Ulcer Scale for Healing (PUSH) tool version 3.0 to measure the surface area (cm^2^) of the lesion calibrated from the ruler. The photographs were thus assessed independently of the local research site in Nigeria and none of the photographs included patient identification or time information – the observers were then blind to intervention status.

### Quantitative outcomes

There were two ulcer outcomes:

1. The total number of ulcers (hands and feet).
2. The area (cm^2^) of the ulcer identified as the largest ulcer at the first (baseline) visit and then subsequently followed up. (Note that this was a protocol deviation – it was originally intended that all ulcers, including any on hands or lower leg, would be followed-up).

Two questionnaires were administered to all participants at all visits in sequence as follows:

a. A general health related Quality of Life questionnaire in the form of the EQ VAS (visual analogue scale): this records the patient’s self-rated health on a vertical 0 to 100 scale from ‘Best imaginable health state’ to ‘Worst imaginable health state’.
b. A life satisfaction (subjective wellbeing) questionnaire (24, 25).This instrument comprises of five questions (all scored out of 10):

i. A cognitive evaluation of the participant’s level of life satisfaction, Question 1 (Q1);
ii. The ‘eudemonic’ concept of whether the things the participant does in their life are worthwhile (Q2); and
iii. Characterise the participant’s affective state on the previous day (Q3, Q4, and Q5).

### Mitigation of bias

Our primary outcomes were number of current ulcers and surface area of largest ulcer at baseline. Observations were made by the unmasked researcher and subsequently compared to masked observations of photographs taken in a standardised manner.

### Statistical Analysis

The statistical methods are fully described in the study protocol; here we provide a brief overview. We used a Bayesian approach to estimation of treatment effects. Analysis of the number of ulcers assumed the outcome to be an autoregressive series of count data within patients, given the strong persistence of ulcers over the time of the study. We specified an integer autoregressive (INAR) model for this outcome, which consists of an autoregressive “thinning” process for existing ulcers and a random Poisson distributed rate of new ulcer formation (26). A logit model was specified for the thinning or persistence of ulcers such that the estimated treatment effect is an odds ratio describing the odds of an ulcer persisting from one period to the next under intervention versus control conditions. The area of the largest ulcer was modelled using a log-normal model conditional on having an ulcer, such that the estimated treatment effect is the relative area of the largest ulcer in intervention and control conditions among those with an ulcer. Wellbeing outcomes were all analysed using a linear model. All models included cluster and cluster-period random effects terms. We used weakly informative prior distributions for model parameters and hyperparameters, which are described in the study protocol.

### Sample size calculation and design analysis

We simulated data from the prior predictive models specified above to evaluate the pre-posterior distributions of treatment effect parameters. We assumed 20 patients per cluster (i.e. per hub) would be recruited. We estimated the average posterior variance, average width of a 95% CrI interval, and minimum effect sizes for which there would be an 80% probability of the 95% CrI excluding zero.

For ulcer count persistence, the smallest effect size for which there is an 80% probability that the 95% credible interval would exclude zero is an odds ratio of 0.3. To translate this into absolute probability terms, given a baseline of 90% probability of persisting, it is equivalent to a reduction of approximately 20 percentage points. For ulcer area, the smallest effect size for which there is an 80% probability that the 95% credible interval would exclude zero is a 42% reduction in largest ulcer size.

### Process evaluation

#### Data Collection

Data were collected from 10 SCGs in 10 villages during the ‘active’ phase of the self-care intervention to assess intervention fidelity/adaptation and to understand how participants experience the intervention. These 10 SCGs were chosen to provide a spread across the different locations. We explored challenges to delivery of the intervention and factors that may facilitate or impede its impact. Data was collected using semi-structured checklists, templates and guides (see Interview Topic Guides and Checklist in Supplementary material, S2) and included:

a. Observations of a sub-set of n=25 intervention sessions from 5 SCGs: Behaviour change techniques used during the group meeting were recorded using a checklist. The researcher observed and recorded how different facilitators/group leaders interpreted their role, how they interacted with the group and how group members responded. These observations helped to understand the fidelity with which the *principles* of the self-care intervention were applied and any local deviations. For 2 SCGs, meetings after the active phase of the intervention were also observed (n=4). Researchers found that these groups had stabilised and there was not much change or any new elements introduced by the groups.
b. Set of notes taken by facilitators summarising the use of the intervention manual and topics of discussion between group members during all 6 initial group meetings for all 10 groups involved in the process evaluation
c. Experiences of the intervention were explored in focus group sessions with 8 self-care groups (8-10 participants each) carried out by the researchers after an observed session; 5 groups had similar numbers of male and female members; 2 were comprised of mostly men and 1 mostly of women. Interviews were conducted with elected leaders of self-help groups (n=3) facilitators of observed sessions (n=3), and key informants i.e. local influential community leaders (n=6) and healthcare professionals (n=6). Participants discussed experiences of self-care interventions and their logistics, and the perceived effect of self-care on group members and the wider community (See Interview Topic Guides and Checklist in Supplementary material, S2). We sampled for diversity of location (sequence) and different perspectives on the intervention. Interviews and focus groups were audio recorded and transcribed.
d. Membership and attendance records were available for 9 out the 10 groups.

#### Qualitative data analysis

We used framework method (27) for the thematic analysis of qualitative data. This method was selected for its suitability in working with teams of multi-disciplinary researchers with varying qualitative analysis experience and large qualitative data. In consultation with all team members, a single matrix for data analysis was developed and utilized in coding transcripts within four key themes:

- Implementation: intervention delivery and what was delivered to participants.
- Context: factors that influenced or were affected by the intervention and its outcomes, and that prevent or enable change prompted by the intervention
- Mechanisms of impact: how participants responded, what mediated this, and any unanticipated pathways and consequences.
- Psycho-social Outcomes: the impact of the intervention on the participants lived experience and that of the community.

Within our frameworks for each of the above, we analysed the data by comparing data across sites and between data sources.

## Results

### Baseline characteristics

Table 1 presents the baseline characteristics of the enrolled participants by trial sequence. The study included 148 unique participants (less than the 240 target).

Among the five sequences, sequence 1 had 39 participants, which contributed the largest proportion of the total number of participants (26%), while sequence 4 had 20 participants contributing 14% of the total. The average age in sequence 1 to 5 were 55.92 (sd 16.15), 54.67 (sd 15.89), 46.47 (sd 18.34), 50.10 (sd 18.04) and 59.03 (sd 18.74), respectively. The gender distribution varied across sequences, with sequences 1 and 5 having more males and sequences 2, 3 and 4 having more females. In terms of education, sequences 1, 4 and 5 had the highest proportion of participants who had never attended school, while sequences 2 and 3 had the most participants who completed primary education. Across all sequences, most participants were self-employed or business owners. In sequence 1, 2 and 3, most participants had a monthly income below 5000 while in sequence 4 and 5, most participants had a monthly income between 5000 and 9999.

The mean duration across all sequences was 58.55 months (sd 94.49), and the median duration was 10 months (IQR 2.25 – 60). However, there was considerable variation in the duration across the five sequences. The overall mean duration was 58.55 months (sd 94.49), however sequence 2 had the shortest mean duration at 16.22 months (sd 18.85), while Sequence 3 had the longest average duration at 95.62 months (sd 135.60).

The overall disability rate was 65.99%. Over half of the participants in sequences 1 (100%), 2 (76%), and 3 (77%) reported having a disability.

### Data completeness

Table 2 summarises the availability of data and missing data patterns for the four observation occasions in the trial (Baseline 1, Baseline 2, Post-treatment 1 and post-treatment 2), in terms of ulcer area. There are 149 unique individuals in the dataset, of which 124 (83%) have the complete set of four observations. One participant’s ulcer (identified whilst carrying out data quality checks) was measured on different limbs across the time periods and was therefore not included in the study. Thus, 148 participants were included for analysis.

**Table 2:** Missing data and follow-up patterns in the data.

|  | Baseline 1 | Baseline 2 | Post-treatment 1 | Post-treatment 2 | n (%) |
| --- | --- | --- | --- | --- | --- |
| Observations<br>(N = missing,<br>Y =<br>observed)) | Y | Y | Y | Y | 124 (83%) |
|  | Y | Y | Y | N | 6 (4%) |
|  | Y | Y | N | Y | 1 (1%) |
|  | Y | Y | N | N | 7 (5%) |
|  | Y | N | Y | Y | 3 (2%) |
|  | Y | N | N | N | 8 (5%) |
| Total |  |  |  |  | 149 (100%) |

### Quantitative Outcomes

The upper plot in Figure 4 shows the baseline (log) ulcer area by trial sequence and observation period. Only observations where an ulcer was present (area > 0 cm2) are included. There was no visual evidence of a decrease in the area of the largest ulcer over the trial. The bottom plot in Figure 4 shows the number of ulcers across four observation periods. Following the intervention, there were small decreases in the proportion of participants with one ulcer (34% and 32% at baseline 1 and 2 decreasing1 to 22% and 21% in post-treatment 1 and 2) or two ulcers (3% and 2.2% in baseline 1 and 2, 1.5% and 1.5% in post-treatment 1 and 2). Correspondingly, there was an increase in the proportion of people with no ulcers (62% and 65% in baseline 1 and 2, 75% and 76% in post-treatment 1 and 2). The change for all outcomes of the trial by observation period for all observations is summarised in Supplementary materials, S3.

**Figure 4:**
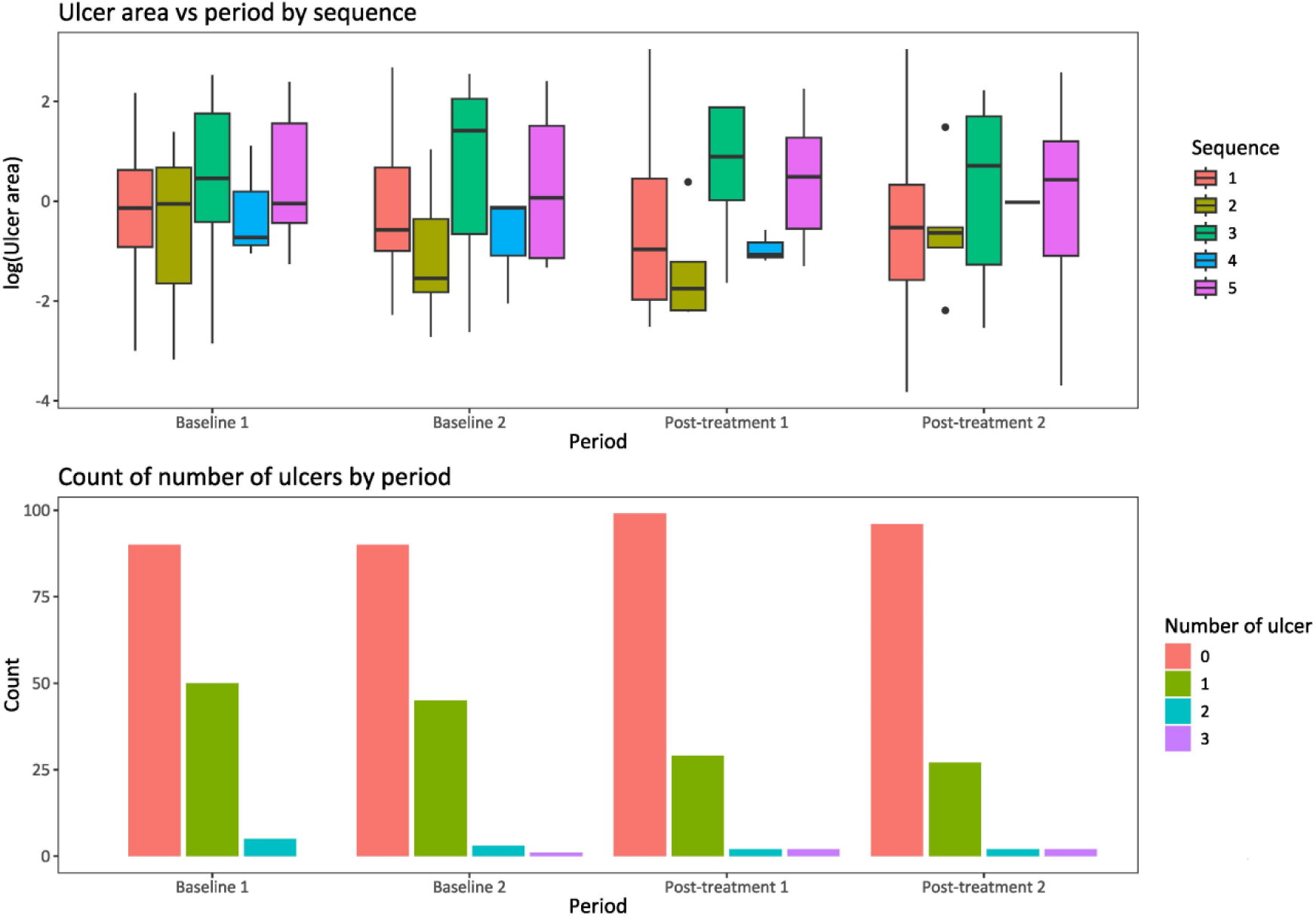
Ulcer area and number of ulcers.

The estimated intervention effects are reported in Table 3. The posterior mean of the relative difference in ulcer area was 1.12 (95% CrI: 0.53, 2.27). The posterior mean odd ratio for ulcer count persistence number of ulcers was 1.00 (95% CrI: 0.29, 3.56). There was thus little difference in ulcer area or number associated with the intervention, although the results were highly uncertain.

**Table 3:**
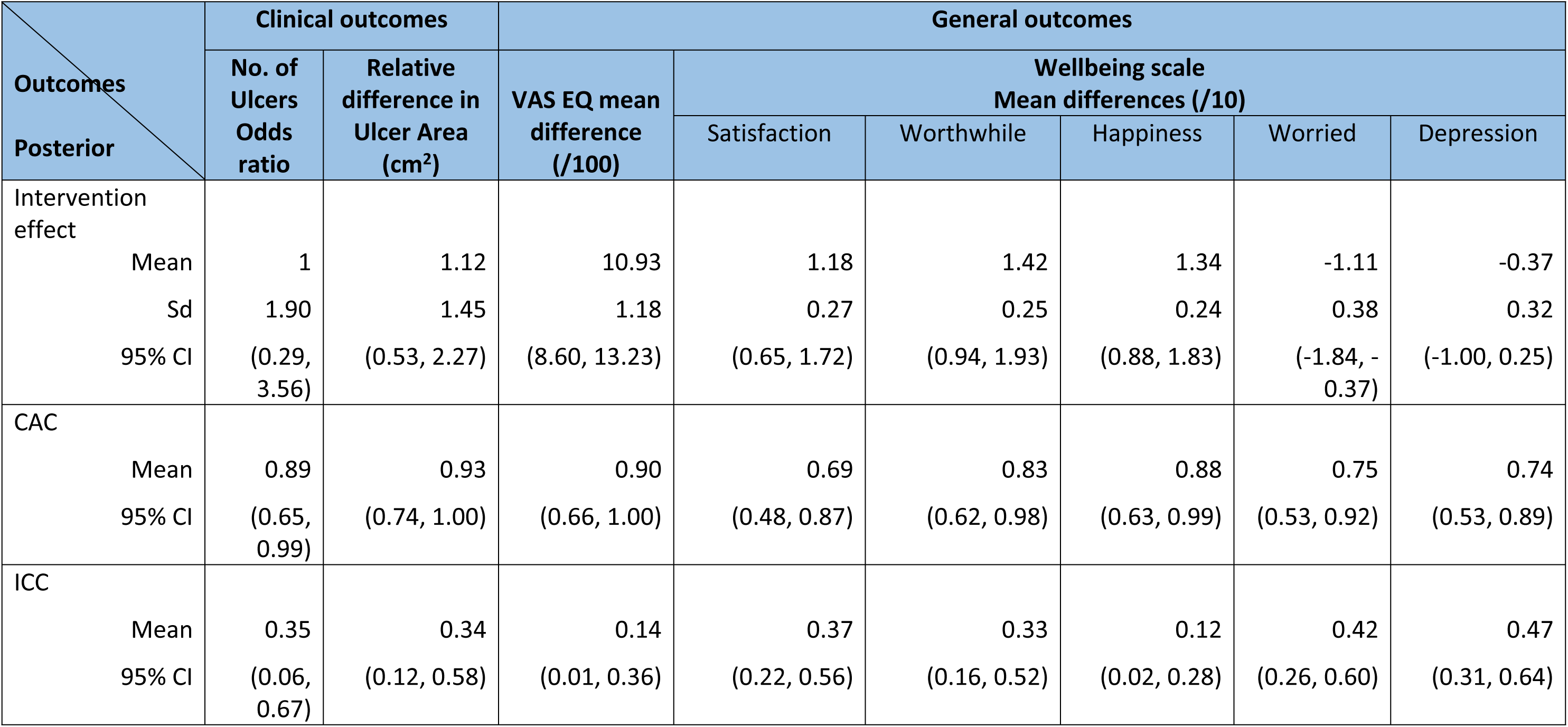
analysis of outcomes.

The psychological outcomes are reported in Table 3. The intervention resulted in an increase of 10.93 (95% CrI: 8.60, 13.23) out of 100 in VAS EQ scores. For the wellbeing scales (ranging from 0 to 10), the satisfaction, worthwhile and happiness scales all showed a positive relationship with the intervention, with increases of 1.18, 1.42, and 1.34 respectively. Conversely, the worried scale decreased by 1.11, and the depression scale decreased by 0.37. These effects are all in the hypothesised direction with confidence limits excluding the null in all cases except depression.

### Process evaluation

Table 4 provides a table of quotes to support the discussed sub-themes for the process evaluation.

**Table 4:**
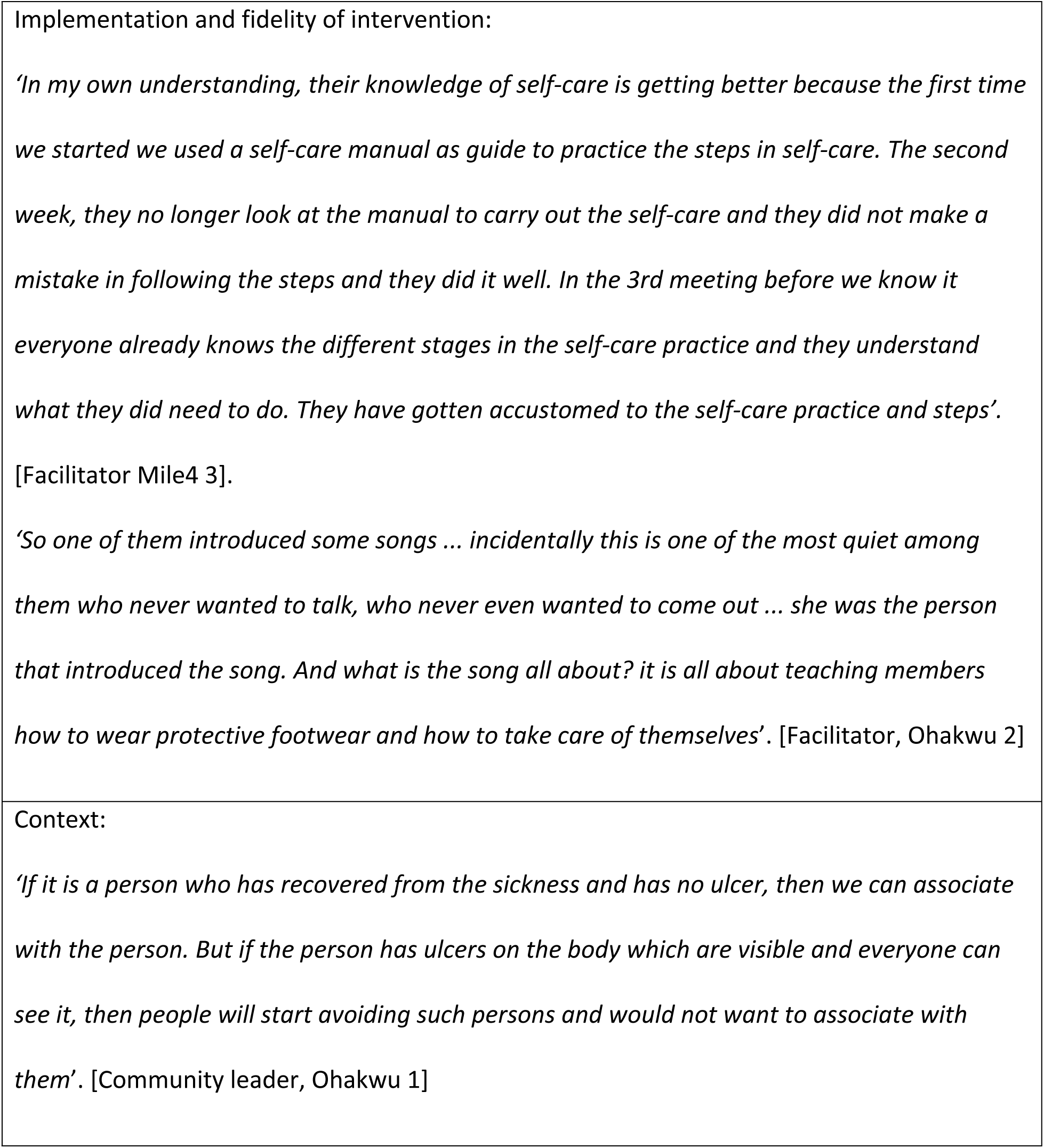
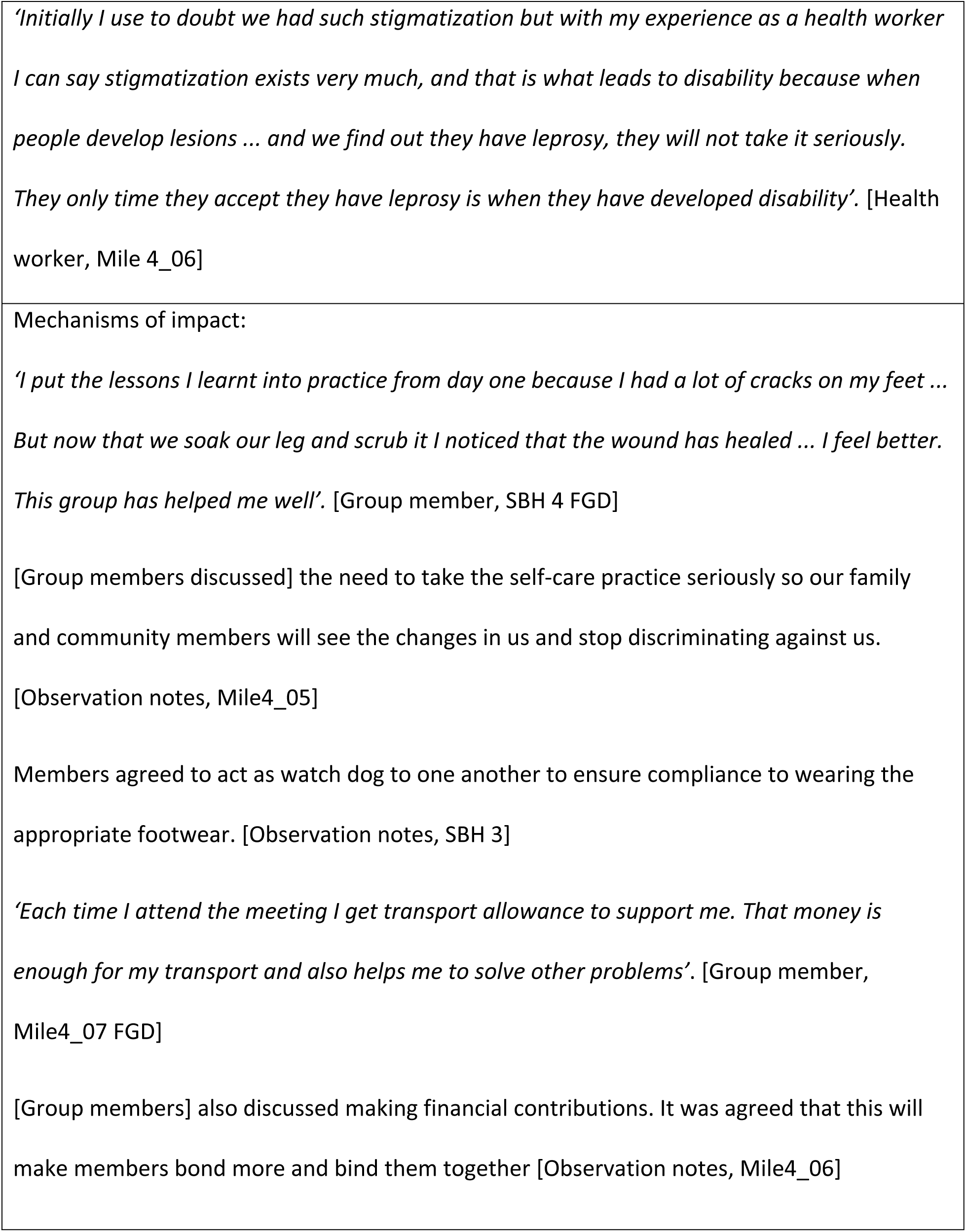
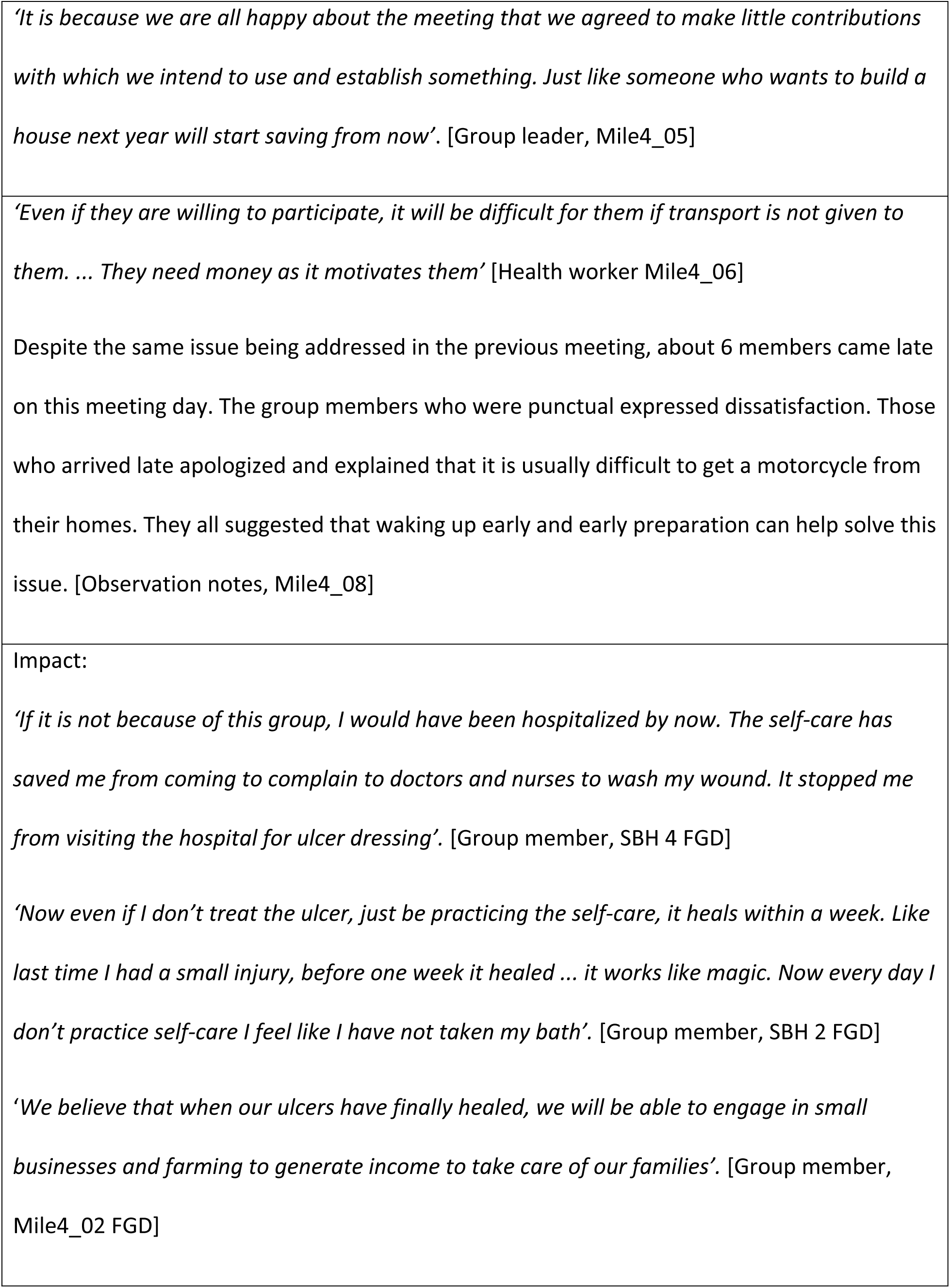
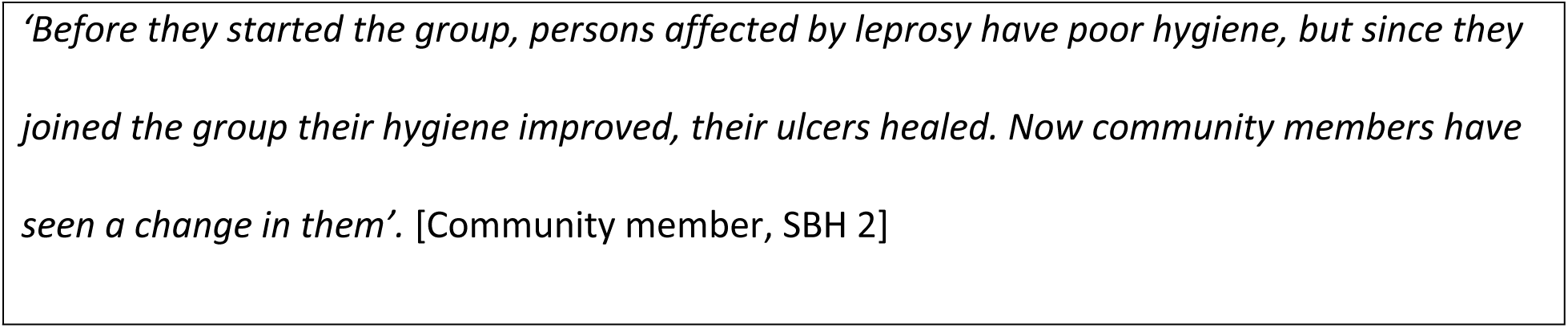
Quotes from the qualitative evaluation.

### Implementation of intervention

As stated, the intervention was implemented in 15 groups of about 10 members in 2 study sites. An average of 8 members attended each meeting during the implementation phase of the intervention and 83% of participants attended all pre and post intervention sessions.

This means that the ‘dose’ of the intervention, i.e. attendance, was fairly consistent; however, there were some contextual factors that might explain variation outlined below. Eight of the 15 groups made plans to continue into the ‘sustainability phase’ at the end of the intervention. Facilitator notes and observer checklists both showed that the manual was used by all groups who settled into a routine and did not need to refer to the manual at the end of six weeks. Observer checklists noted that a range of techniques was used by group leaders and facilitators e.g. raising positive expectations of ulcer improvement with self-care and encouraging role modelling for other members of the community. Group members were also aware that self-care had to be continued at home. In 5 groups, leaders made adaptations to the format by asking group members to take turns to lead activities which made meetings more inclusive. One group developed a song to help them remember the steps of self-care outlined in the manual.

### Context

The main contextual factors we identified were work patterns, availability of care and stigma for people with leprosy. Work patterns were important as in some areas farming was small scale and did not impact attendance at meetings whereas in others producing crops for sale and attending markets could impact attendance. Some group members also discussed needing to ride a motorbike for work which could contribute to the development of ulcers. While leprosy care is available from the participating hospital sites, some focus group participants said they had used traditional and faith-based healers as their first port of call, due to proximity and a general belief that leprosy is caused by witchcraft. Stigma for those affected by leprosy was widespread with interviewees discussing being shunned by former friends and being unable to do business or trade farm produce.

### Mechanisms of impact

Overcoming stigma was a main motivator to persist with self-care. Many group members referred to provision of fresh bandages, which they felt had reduced smell from ulcers, improved their standing in the community and enhanced their ability to trade. Another motivator for many was noticing change in wound healing and cracked skin; some also anticipated being completely healed in the future. The provision of incentives to group members played a role, with fresh bandages contributing to the neat look outlined above, and money to help with transport, some of which could be used to solve other problems. Group members identified the advantages of group cohesion and mutual support. A striking finding was that seven out the 10 groups we observed had transformed themselves into self-help groups by forming a savings club to help with the cost of self-care or to support group members more widely. There were also many examples of group members being supportive to each other: applauding each other for good results; giving advice on improving self-care; or counselling people who developed ulcers on how to avoid them in the future. In four groups, members agreed to watch each other outside of the group and call out any lapses, e.g. not using protective footwear.

Challenges and barriers were mostly around transport as in one of the two sites some of the group members lived at large distances from the meeting venue. In these situations, the availability and cost of motorcycle taxis led to a few members missing groups or arriving late for meetings. Sustainability of the groups was another concern as members anticipated difficulties when funding for the self-care materials and transport was discontinued. Taking part in group activities was more challenging for people who were elderly or spoke a different language from other group members; however, observation notes indicate that younger people would step in and help with translation.

### Impact

The perceived impact of the intervention on the participants’ lived experience centred around gaining confidence to practice self-care; some believed that their health would have been much worse without the group. As outlined above, they also saw another important impact in improved hygiene and appearance and anticipated that their lives would continue to get better as they would be able to sell farm produce or return to work. This was echoed by members of the community we interviewed. Group members and health care workers also said that there now was less need to go to the hospital for treatment as they improved with self-care.

## Discussion

### No improvement in ulcers

The main motivation for this study was to reduce the ulcer ‘burden’ in terms of the number and size of ulcers. We convened an international expert group to develop guidelines, and the guidelines covered not just the clinical aspects, but included implementation science principles covering the system as a whole. RedAid Nigeria was committed to this project and devoted resources and effort into making it a success. Yet, there was no evidence for an improvement in ulcer status among participants. That said, the results were highly uncertain, partly reflecting the under-recruitment of patients into the trial. For example, mean difference in ulcer area was compatible with approximately a halving (or doubling) in ulcer area. That said our point estimates are close to the null value and exclude the large effect (halving of ulcer areas) that might have been anticipated.

### Impact of the intervention

The lack of effect we observed occurred despite the observation that the intervention was implemented as intended, sessions were well attended and were well received by study participants. In line with this, six measures of health-related quality of life and well-being improved. The results crossed the line 95% probability level in five of these. The effect sizes are material – for example, four of the five well-being scales improved by ten points on a 100-point scale. The fact that different clusters received the intervention at different calendar times suggests that there was not a seasonal effect. The qualitative data suggest that participants were able to continue with their normal lives (an aim of self-care) more confidently and successfully than before the intervention. Their ability to present themselves more neatly and without odour from ulcers was important for their livelihoods and reduced their perceived stigma (28, 29). Self-help group members gained in confidence through the psychological and financial support of self-help group membership to manage their ulcers well. The qualitative data show that participants and others believed that their ulcers were improving. This is consistent with the observed psycho-therapeutic benefit of the intervention but not with the objective ulcer measurement. The participants innovated using song and graduated from self-care to self-help by forming savings syndicates – a phenomenon also encountered in the famous women’s groups studies (30). In short, the intervention was delivered as intended; it was appreciated and taken up enthusiastically and it produced substantial gain in wellbeing.

### The nature of the ulcers in the index population

Some clue to the reason for our null findings on ulcer healing may lie in the population of people participating. Although economically active, they were an elderly group (well over 50 on average), where most had very chronic ulcers and the rate of deformity was also high reaching 100% in one of our sequences (Table 1). In short, this was a population with what might be termed ‘indolent’ ulcers associated with extensive scarring and deformity, both of which effect ulcer healing (31–33). In short, we think that a plausible reason for our findings is that “it was too little, too late”. If this is correct, then our findings have two corollaries: -

1. They emphasise the importance of preventing ulcers from forming and intensive early care when they do – techniques such as off-loading devices (4, 5) and re-innervation (34) may be extremely cost-effective in this regard.
2. Interventions for indolent ulcers might have to be more intensive, say at admission and / or plaster casting to ensure satisfactory off-loading.

### Methodological aspects / strengths and weaknesses

Our study was designed to have high validity involving randomisation and blinding outcome assessment where possible. However, it is one study in one context. Therefore, we do not think our findings should be generalised across all community leprosy care. Given what we have said regarding the ‘population’ participating in our study– our findings are not directly referrable to a setting where people have ulcers of considerably shorter duration, where those ulcers were smaller, and where deformity is the exception. However, many places where ulcers are endemic have large numbers of people with chronic ulcers living in extreme poverty. Our study site is emblematic of many other circumstances where leprosy ulcers occur.

Our study has further limitations; the number of participants was lower than intended and hence our observations of ulcer size and number are imprecise. The duration of the study period was limited by the duration of the grant (five years) during which we had to update guidelines, adapt the intervention, recruit participants, collect baseline readings, roll out the intervention sequentially, obtain independent ulcer measurements and complete the analysis and write-up. We have noted elsewhere that the typical funding cycle preludes longer-term observations of impact and sustainability.

## Data Availability

10.6084/m9.figshare.28311938

https://figshare.com/account/items/28311938

## DECLARATIONS

### Ethics approval and consent to participate

This research has been performed in accordance with the Declaration of Helsinki for Human Research of the World Medical Association. Approval was granted by the University of Birmingham Biomedical and Scientific Research Ethics Committee (BSREC) and locally in Nigeria through the University of Nigeria Teaching Hospital Health Research Ethics Committee (NHREC 0501-2008B).

Eligible participants were given a Participant Information Sheet in the local language. For non-literate participants, information was provided verbally. Written informed consent was obtained from all willing to participate; or thumb/fingerprints was requested in lieu of a signature where necessary.

### Competing interests

The authors have no competing interests.

### Funding

This research was funded by the National Institute for Health and Care Research (NIHR: 200132) using UK Aid from the UK Government to support global health research. RJL is also funded by NIHR Applied Research Collaboration (ARC) West Midlands. RJL, YW, SC and JS are funded by NIHR Midlands Patient Safety Research Collaboration (PSRC). The views expressed in this publication are those of the author(s) and not necessarily those of the NIHR or the UK Department of Health and Social Care.

### Authors’ contributions

JS, JC, NO, NN, SC, LU, UA, and OI: contributed to the development, drafting, and editing of the protocol and paper.

SIW: contributed to the evaluation design, sample size calculation, randomization, quantitative sample selection, and critically reviewed the intellectual content of both the protocol and paper.

YW and AL: contributed to data analysis and the drafting and editing of the paper. PG and FG: contributed to the development and critical review of the intellectual content of the protocol and paper.

AL and FG: contributed to the development, drafting, and editing of the paper. SC: helped draft the protocol and paper, managed data flow between the data collection site and blinded assessors, organized staff training on ulcer measurement, ensured GCP compliance, and coordinated the steering committee.

AT: As Principal Investigator, contributed to the study’s conception, protocol development, critically reviewed the intellectual content, and drafted the initial version of the paper. RJL: Principal Investigator on the NIHR grant (below), contributed to the study’s conception, co-designed the evaluation, critically reviewed the intellectual content, and assisted in drafting the paper.

All authors reviewed and approved the final manuscript.

## Acknowledgements

We extend our gratitude to the patients and trial team members at the recruitment site for their contributions to data collection and entry. We want to thank Anjali Shrivastva and Khushboo Johri from The Leprosy Mission Trust India, who performed blinded assessments of all photographs of ulcers.

## Notes

### Competing Interest Statement

The authors have declared no competing interest.

### Clinical Trial

ISRCTN12572449

### Clinical Protocols

https://osf.io/u2r5s

### Author Declarations

1. Locally in Nigeria through the University of Nigeria Teaching Hospital Health Research Ethics Committee (NHREC 0501-2008B). 2. University of Birmingham Biomedical and Scientific Research Ethics Committee (BSREC)

